# Hospital and operator procedural volumes and one-year outcomes for TAVR in the United States: A STS/ACC TVT Registry analysis

**DOI:** 10.64898/2026.07.13.26358001

**Authors:** Dharam J. Kumbhani, Wayne Batchelor, Joseph Cleveland, Pratik Manandhar, Andrzej Kosinski, Samir Kapadia, Gorav Ailawadi, Gregory Fontana, Andrei M. Pop, Saket Girotra, James A. de Lemos, John Carroll, Ralph Brindis, Tsuyoshi Kaneko, Vinod H. Thourani, Robert W Yeh, Amit N. Vora, Michael Mack, Vinay Badhwar, Roxana Mehran, Sreekanth Vemulapalli

## Abstract

**Background:** Prior analyses have demonstrated an inverse association between transcatheter aortic valve replacement (TAVR) procedural volume and short-term outcomes. However, less is known regarding the relationship between procedural volume and 1-year outcomes in the contemporary TAVR era.

**Objectives:** To evaluate the association between annual hospital and operator TAVR procedural volumes and 1-year clinical outcomes in a contemporary national cohort.

**Methods:** Clinical records from the Society of Thoracic Surgeons (STS)/American College of Cardiology (ACC) Transcatheter Valve Therapies (TVT) Registry for patients undergoing commercial TAVR between January 2020 and December 2022 were linked to Centers for Medicare & Medicaid Services administrative claims. Annualized hospital and operator TAVR volumes were modeled continuously and categorized into tertiles. Primary outcomes included 1-year all-cause mortality, stroke, the composite of mortality or stroke, and all-cause readmissions. Hierarchical risk-adjusted models accounting for patient clustering within sites were used to evaluate associations between procedural volume and outcomes.

**Results:** Among 215,335 patients undergoing TAVR at 788 hospitals by 3,444 operators between 2020 and 2022, median annual hospital and operator volumes were 74 (IQR: 43-115) and 16 (IQR: 10-32), respectively. Volume was then categorized into tertiles (low, medium and high). Compared with high-volume hospitals (≥102/year), low-volume hospitals (≤52/year) had higher adjusted rates of 1-year all-cause mortality (Odds Ratio (OR): 1.10 [95% CI: 1.05-1.16]), stroke (OR: 1.10 [95% CI: 1.01-1.19]), mortality or stroke (OR: 1.10 [95% CI: 1.05-1.15]), and all-cause readmissions (OR: 1.05 [95% CI: 1.00-1.09]). Compared with high-volume operators (≥25/year), low-volume operators (≤11/year) had higher adjusted rates of stroke (OR: 1.16 [95% CI: 1.05-1.28]) and mortality or stroke (OR: 1.09 [95% CI: 1.03-1.15]) but not other endpoints.

**Conclusions:** In a large, contemporary national TAVR registry, lower annual hospital (≤ 52/year) and operator (≤ 11/year) procedural volumes were independently associated with worse 1-year clinical outcomes. These findings suggest that procedural experience continues to influence outcomes despite maturation of contemporary TAVR practice.

Central illustration:
Central illustration

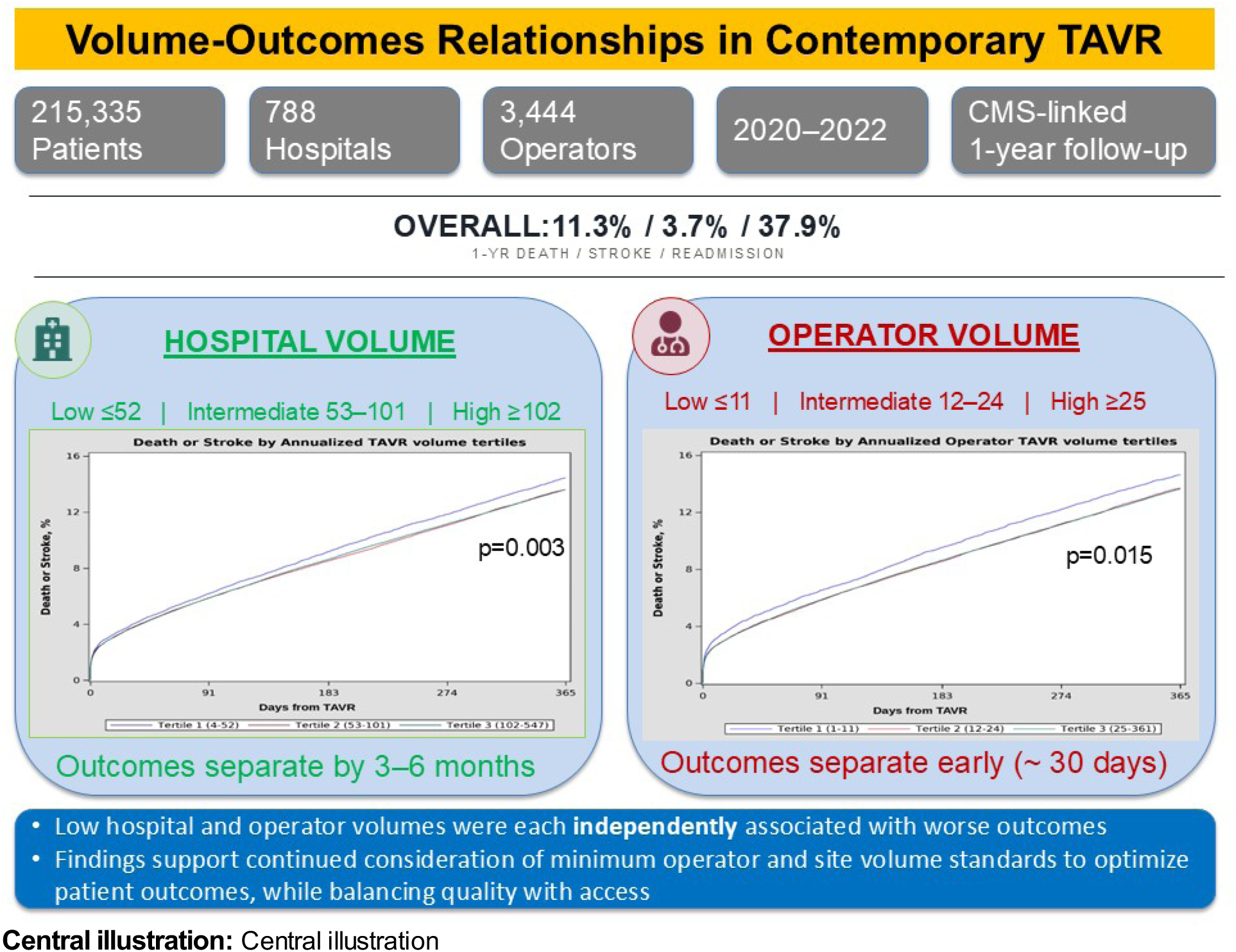

## Introduction

Aortic stenosis (AS) remains one of the most common valvular heart diseases worldwide, and transcatheter aortic valve replacement (TAVR) has become the predominant treatment strategy for patients with severe symptomatic AS.(1–3) The rapid expansion of TAVR programs and operators across the United States has been accompanied by longstanding interest in procedural volume as a marker of quality and a basis for regulatory and credentialing requirements.(4–6) Prior studies have demonstrated inverse associations between hospital and operator TAVR volumes and clinical outcomes. Carroll and colleagues evaluated 42,988 high- and extreme-risk patients undergoing TAVR at 395 hospitals between 2011 and 2015 and observed a strong volume-outcome relationship, with higher hospital TAVR volumes associated with lower in-hospital mortality.(7) In a subsequent analysis of intermediate and high/extreme risk patients undergoing TAVR between 2015 and 2017 at 555 hospitals, risk-adjusted 30-day mortality was higher in the lowest-volume quartile for both hospitals and operators.(8) In two subsequent analyses in the low-risk era, a persistent inverse association between hospital as well as operator volumes and 30-day outcomes was observed.(9,10) However, with increasing operator experience, maturation of TAVR technology, and expansion to lower-risk populations, the volume–outcomes relationship has progressively flattened, particularly for short-term mortality. However, its association with longer-term outcomes, including 1-year mortality, stroke, and readmissions, remains less well defined. This is particularly relevant as TAVR is increasingly performed in lower-risk patients with longer life expectancy. Accordingly, we evaluated the association between hospital and operator volume and 1-year outcomes in a large contemporary national cohort undergoing TAVR.

## Methods

### Data source

Details of the Society of Thoracic Surgeons (STS) /American College of Cardiology (ACC) Transcatheter Valve Therapies (TVT) Registry (R) have been published previously.(11) As reported, this registry, launched in 2011, captures data on all U.S. patients undergoing TAVR with commercially approved devices, fulfilling the CMS requirement for data submission to a national registry. Data integrity is supported through standardized data elements, National Cardiovascular Data Registry (NCDR) quality checks, and annual independent audits. Use of TVT-R data for research was approved by institutional review boards at Advarra and Duke Clinical Research Institute (DCRI), with a waiver of informed consent.

### Study population

TVT-R clinical records for TAVR procedures performed between January 2020 and December 2022 were linked to Medicare administrative claims (Medicare Advantage [MA] and Medicare Fee For Service [FFS]). Linkage was achieved using a deterministic multi-rule algorithm based on indirect identifiers (date of birth, sex, admission and discharge dates, and hospital site) previously validated for cardiovascular registry linkage.(12) Claims filed through the end of December 2023 were included in the analysis set to allow for at least 1 year follow-up for all patients. Patient records from hospitals that had performed < 20 TAVR procedures over the study period (n=289), or with < 1 year of data in the TVT registry (n=41) or that could not be linked to Medicare claims data (n=26,823) were excluded (**Supplemental Figure**).

### Calculation of Hospital and Operator Volumes

Annualized TAVR volume was defined as 12 times the mean monthly number of TAVR procedures performed over the interval between the first and last procedure months.

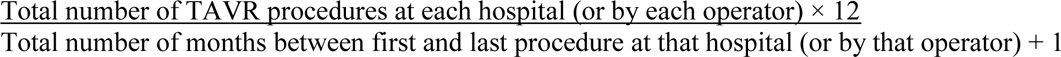

The total number of TAVR procedures performed by each operator was determined using their unique NPI, allowing accurate tracking across hospitals, and as previously reported.(8,10) Since CMS requires at least two operators (an interventional cardiologist and a cardiac surgeon) for TAVR, we assigned the operator with the highest lifetime volume (calculated from inception of the TVT registry to December 2022) as primary.(10)

### Study Outcomes

The primary outcomes were all cause mortality, stroke, the composite of all cause mortality or stroke, all cause readmission, and home time at 1 year after the index TAVR procedure. These outcomes were compared across low, intermediate, and high annual hospital and operator TAVR volume tertiles. Patient follow-up was considered to be censored at the end of the follow-up period (December 31, 2023). Linked CMS claims files were used to create a longitudinal record including vital status and rehospitalization events, using validated techniques.(12–14) Following hospital discharge, death was identified using the Medicare Denominator file. Stroke was identified from Medicare inpatient claims using ICD-10-CM diagnosis codes I60x, I61x, I62x, I63x, I6781, I6782, I6789, I679, I97810, I97811, I97820, I97821, G9731, G9732, consistent with previously validated claims-based algorithms.(15)

### Statistical analysis

The hospital or operator was the unit of analysis for assignment to a volume group. However, the patient was the unit of analysis for clinical variables and outcomes. Hospital and operator specific TAVR volume was first calculated, ordered and categorized such that each tertile (low, intermediate, high) had the approximately the same number of sites or operators.

Patient demographics, baseline characteristics and procedure characteristics were compared across hospital and operator annualized TAVR volume and tested for association. Categorical variables were presented as frequencies and assessed using Pearson chi-square test, and continuous variables were presented as median (q1,q3) and assessed using Kruskal-Wallis test. Cumulative rates for death, stroke, composite of death or stroke, all-cause hospital readmissions and home time for 365 day period are also provided by volume tertiles.

Volume-Outcomes Analyses: Adjusted rates for binary outcomes of interest were estimated using a generalized linear mixed model with binomial distribution and logit link, with risk adjustment based on variables specified in **Supplemental Methods.** The 2-level hierarchical structure of the data with patients nested within sites/operators and a random residual was used to account for clustering within sites/operators. To mitigate non-convergence commonly encountered in hierarchical models with multiple covariates, a single composite risk score was used in place of individual adjustment variables. This risk score was derived from a logistic regression model estimating the log-odds of each outcome and subsequently incorporated as a single covariate in the adjusted hierarchical models. For the outcome of 1-year home time, a Poisson generalized linear mixed model with a log link was used to evaluate the association with volume tertiles. The 2-level hierarchical structure of the data with patients nested within operators and random residuals was used to account for clustering within sites/operators. Finally, cubic spline transformation of parameter estimates was used to visualize relationships between TAVR volume and outcomes. Number needed to harm (NNH) was calculated as the inverse of the absolute difference in adjusted event rates between low- and high-volume tertiles. For the stroke volume-outcome analyses, patients who died within 1 year without a stroke were excluded from the analysis. Home-time was calculated as the number of days within 365 days after the TAVR procedure date spent alive at home. The day of TAVR was considered day 0, and then for each patient, we calculated the number of days spent in a facility (SNF, LTAC, or an acute care hospital) within the first 365 days and subtracted them from the 1-year home time. If a patient died within 365 days from the procedure date, days lost because of death were also subtracted from home (16).

We also evaluated the interaction between operator and hospital volumes. To minimize chance findings from multiple comparisons, we predefined interaction testing primarily to assess whether a low volume operator at a high volume site or vice versa performs differently. Accordingly, the following interaction tests were pursued: high-volume operator/high-volume hospital, low-volume operator/high-volume hospital, and low-volume operator/low-volume hospital.

For missing adjustment variables, simple imputation techniques were used: continuous variables were median imputed and categorical variables were imputed to mode values. Other than information regarding the presence of left main stenosis ≥ 50% (missing: 5.4%) and proximal LAD stenosis ≥ 70% (missing: 5.6%), all other variables had <1% missing information (**Supplemental Methods**).

All statistical analyses were performed at DCRI using SAS version 9.4 (SAS Institute, Cary, NC). All *p*-values were two-tailed, with statistical significance set at 0.05. All confidence intervals (CI) were calculated at the 95 percent level.

## Results

Between January 2020 and December 2022, 215,335 TVT-R records linked to CMS administrative claims data from 788 hospitals and 3,444 operators were identified and included in the final study cohort. **Figure 1** shows the distribution of annual hospital and operator volumes. The median annual hospital and operator TAVR volumes were 74 (IQR: 43-115) and 16 (IQR: 10-32) patients, respectively. Baseline characteristics across the tertiles of annual hospital TAVR volumes are listed in **Table 1**. The median patient age was 80 years, and 44.2% of patients were women. The median STS PROM was 3.8%, and 27.3% were site-reported as low-risk patients. Of the TAVR procedures, 96.1 were done via transfemoral access, 6% were valve-in-valve, and cerebral embolic protection was utilized in 13.8% of patients. Patients undergoing TAVR at low-volume hospitals and by low-volume operators were more likely to have fewer comorbidities (for instance, PAD, prior stroke/TIA, atrial arrhythmias) and were lower-risk patients. Importantly, patients undergoing TAVR at low-volume hospitals and by low-volume operators were more likely to receive general anesthesia, and had longer procedure times, higher contrast use, and greater paravalvular leak and mean gradients post-TAVR.

**Figure 1:**
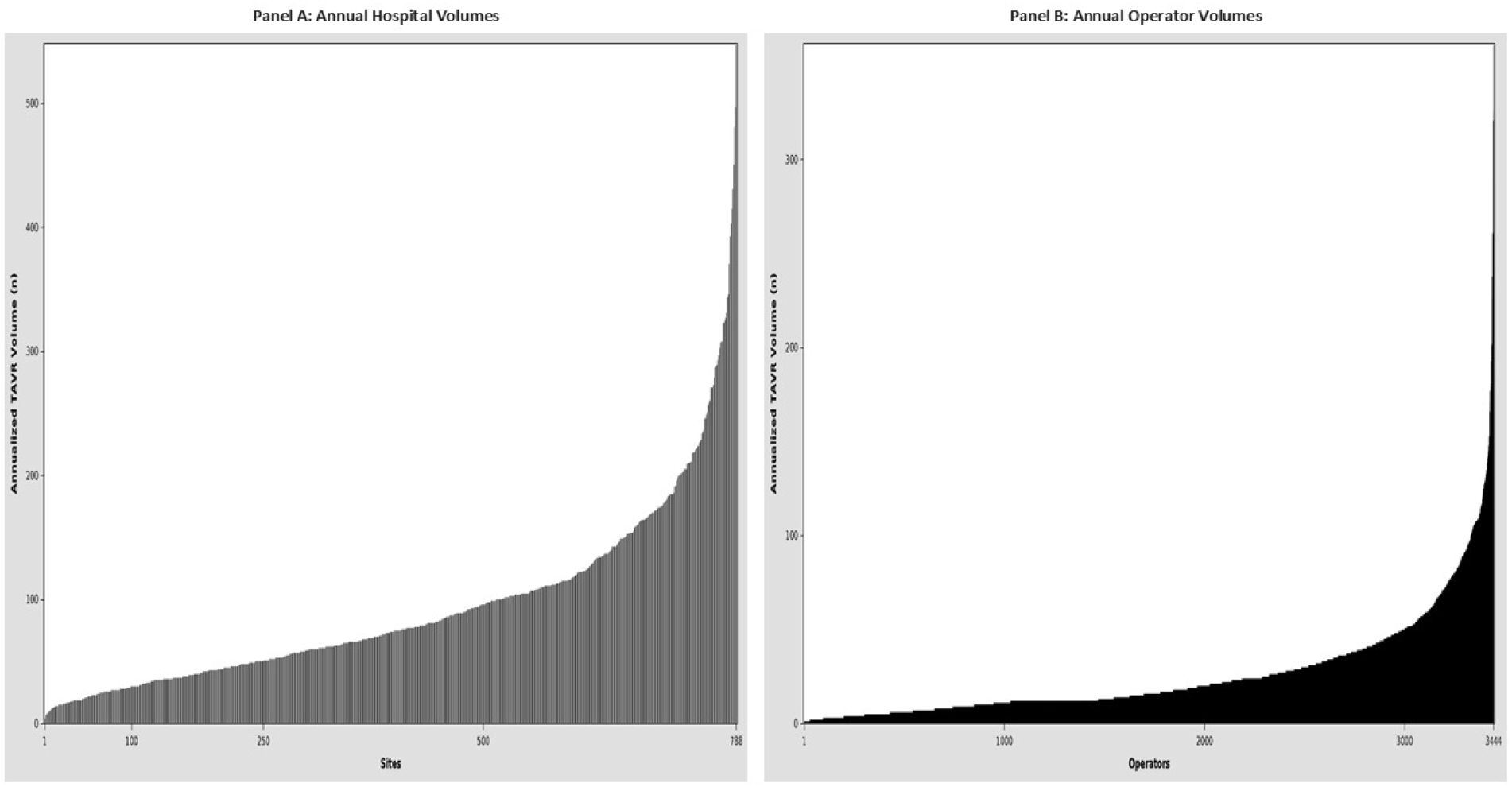
Distribution of annual hospital (panel A) and operator (panel B) volumes for TAVR.

**Table 1.** Patient and clinical characteristics by annualized hospital TAVR volume tertiles.

|  | **Tertile 1**<br>(≤ 52/year)<br>(N =<br>23,645;<br>Hospitals =<br>263) | **Tertile 2**<br>(53-101/year)<br>(N =<br>56,791;<br>Hospitals =<br>261) | **Tertile 3**<br>(≥102/year)<br>(N =<br>134,899;<br>Hospitals =<br>264) |  | **Tertile 1**<br>(≤<br>11/year)<br>(N =<br>11,822;<br>Operators<br>= 1,030) | **Tertile 2**<br>(12-24/year)<br>(N =<br>32,000;<br>Operators<br>= 1,256) | **Tertile 3**<br>(≥25/year)<br>(N =<br>171,113;<br>Operators<br>= 1,158) | **p-**<br>**value** |
| --- | --- | --- | --- | --- | --- | --- | --- | --- |
| *Sociodemographic* |  |  |  |  |  |  |  |  |
| Age (Median, IQR) | 79.0 (74.0,<br>85.0) | 80.0 (74.0,<br>85.0) | 80.0 (74.0,<br>85.0) | <0.001 | 80.0 (74.0,<br>85.0) | 79.0 (74.0,<br>85.0) | 80.0 (74.0,<br>85.0) | <0.001 |
| Female gender, % | 46.7 | 43.9 | 44.0 | <0.001 | 44.9 | 44.4 | 44.1 | 0.14 |
| Non-White non-Hispanic,<br>% | 13.3 | 10.5 | 8.7 | <0.001 | 13.8 | 10.5 | 9.2 | <0.001 |
| Body Surface Area<br>(Median, IQR) | 1.9 ( 1.7,<br>2.1) | 1.9 ( 1.8,<br>2.1) | 1.9 ( 1.7,<br>2.1) | <0.001 | 1.9 ( 1.7,<br>2.1) | 1.9 ( 1.8,<br>2.1) | 1.9 ( 1.7,<br>2.1) | <0.001 |
| *Past medical history* |  |  |  |  |  |  |  |  |
| Diabetes, % | 37.4 | 37.2 | 36.8 | 0.11 | 38.5 | 38.0 | 36.7 | <0.001 |
| Peripheral artery disease,<br>% | 17.5 | 18.4 | 20.7 | <0.001 | 18.1 | 18.1 | 20.2 | <0.001 |
| Current smoker, % | 5.9 | 5.7 | 5.1 | <0.001 | 5.5 | 5.6 | 5.3 | 0.022 |
| Dialysis, % | 3.2 | 3.0 | 2.9 | *0.12* | 3.4 | 3.0 | 3.0 | *0.05* |
| Prior MI, % | 15.2 | 15.8 | 17.3 | *<0.001* | 15.8 | 16.2 | 16.8 | *<0.001* |
| Prior PCI, % | 31.1 | 31.0 | 29.5 | *<0.001* | 30.1 | 30.8 | 29.9 | *0.011* |
| Prior CABG, % | 14.0 | 14.6 | 14.6 | *0.054* | 13.4 | 14.6 | 14.6 | *0.002* |
| Prior cardiac surgery, % | 17.3 | 18.7 | 19.5 | *<0.001* | 17.5 | 18.6 | 19.2 | *<0.001* |
| Prior stroke/TIA, % | 13.9 | 14.9 | 15.9 | *<0.001* | 14.6 | 15.0 | 15.5 | *0.003* |
| Severe lung disease on home oxygen, % | 6.9 | 6.4 | 5.9 | *<0.001* | 4.8 | 5.0 | 4.7 | *0.11* |
| Hostile chest, % | 2.6 | 3.5 | 3.2 | *<0.001* | 3.1 | 2.7 | 3.3 | *<0.001* |
| Porcelain aorta, % | 0.9 | 0.8 | 0.8 | *0.12* | 0.9 | 0.9 | 0.9 | *0.6* |
| Atrial fibrillation or flutter, % | 33.5 | 34.2 | 36.5 | *<0.001* | 34.3 | 35.2 | 35.7 | *0.004* |
| Permanent pacemaker, % | 11.7 | 11.5 | 11.3 | *0.16* | 11.4 | 11.6 | 11.4 | *0.58* |
| Previous ICD, % | 2.3 | 2.5 | 2.6 | *0.043* | 2.8 | 2.4 | 2.5 | *0.099* |
| Conduction defect, % | 27.8 | 30.1 | 35.9 | *<0.001* | 29.3 | 30.4 | 34.4 | *<0.001* |
| NYHA class IV within 2 weeks, % | 8.8 | 9.0 | 8.7 | *0.092* | 8.9 | 8.4 | 8.9 | *0.011* |
| Prior aortic valve procedure, % | 5.9 | 6.8 | 8.7 | *<0.001* | 6.7 | 6.9 | 8.1 | *<0.001* |

| *Laboratory values and testing* |  |  |  |  |  |  |  |  |
| --- | --- | --- | --- | --- | --- | --- | --- | --- |
| LVEF, % (median, IQR) | 60.0 (54.0, 63.0) | 60.0 (53.0, 64.0) | 60.0 (53.0, 65.0) | <0.001 | 60.0 (53.0, 63.0) | 60.0 (53.0, 63.0) | 60.0 (54.0, 65.0) | <0.001 |
| Hemoglobin, g/dL (Median, IQR) | 12.5 (11.1, 13.7) | 12.6 (11.2, 13.8) | 12.5 (11.1, 13.7) | <0.001 | 12.5 (11.1, 13.7) | 12.5 (11.2, 13.7) | 12.5 (11.2, 13.7) | 0.012 |
| eGFR (Median, IQR) | 61.8 (47.4, 78.1) | 61.8 (47.4, 77.9) | 61.9 (47.2, 77.8) | 0.76 | 61.7 (47.2, 77.8) | 61.4 (47.1, 77.1) | 62.0 (47.3, 77.9) | 0.016 |
| Left main stenosis ≥ 50%, % | 5.4 | 5.1 | 5.2 | 0.04 | 5.0 | 5.4 | 5.2 | 0.36 |
| Proximal LAD ≥ 70%, % | 12.1 | 11.9 | 10.8 | <0.001 | 11.2 | 12.2 | 11.1 | <0.001 |
| KCCQ overall score (Median, IQR) | 50.0 (31.3, 69.8) | 50.0 (31.3, 70.8) | 51.0 (31.8, 72.9) | <0.001 | 49.0 (30.2, 68.8) | 50.0 (31.3, 70.8) | 51.0 (31.8, 71.9) | <0.001 |
| Bicuspid aortic valve, % | 9.6 | 9.9 | 9.5 | 0.027 | 10.1 | 9.5 | 9.6 | 0.21 |
| Moderate/severe MR, % | 18.3 | 18.6 | 19.4 | <0.001 | 19.0 | 18.7 | 19.2 | 0.11 |
| Moderate/severe TR, % | 16.0 | 16.1 | 16.4 | 0.14 | 16.2 | 16.3 | 16.3 | 0.97 |
| STS PROM (Median, IQR)* | 3.6 ( 2.2, 5.9) | 3.7 ( 2.3, 6.0) | 3.8 ( 2.4, 6.4) | <0.001 | 3.7 ( 2.3, 6.1) | 3.7 ( 2.3, 6.0) | 3.8 ( 2.4, 6.3) | <0.001 |
| High/extreme risk (site reported), % | 32.5 | 31.2 | 40.3 | <0.001 | 34.3 | 35.0 | 37.6 | <0.001 |
| Low risk (site reported), | 30.6 | 32.8 | 24.5 | <0.001 | 29.9 | 28.9 | 26.9 | <0.001 |
| % |  |  |  |  |  |  |  |  |
| *Procedural characteristics* |  |  |  |  |  |  |  |  |
| Transfemoral access (%) | 97.0 | 96.2 | 95.9 | <0.001 | 96.4 | 96.6 | 96.0 | <0.001 |
| Valve-in-valve procedure (%) | 4.6 | 5.1 | 6.7 | <0.001 | 5.3 | 5.2 | 6.2 | <0.001 |
| Embolic protection use (%) | 8.2 | 9.1 | 16.8 | <0.001 | 10.5 | 10.3 | 14.7 | <0.001 |
| Minimal or moderate/conscious sedation (%) | 48.6 | 51.0 | 63.5 | <0.001 | 54.7 | 55.0 | 59.7 | <0.001 |
| Procedure duration (mins) | 72.0 (55.0, 96.0) | 65.0 (49.0, 86.0) | 62.0 (46.0, 84.0) | <0.001 | 72.0 (55.0, 96.0) | 68.0 (52.0, 90.0) | 63.0 (47.0, 85.0) | <0.001 |
| Contrast volume (ml) | 90.0 (60.0, 126) | 80.0 (55.0, 115) | 70.0 (47.0, 100) | <0.001 | 85.0 (60.0, 123) | 80.0 (54.0, 114) | 74.0 (50.0, 105) | <0.001 |
| Mean gradient at end of procedure or hospital discharge (mean and median values; mmHg) | 10.0 (7.0, 14.0) | 10.0 (7.0, 14.0) | 10.0 (7.0, 13.0) | <0.001 | 10.0 (7.0, 14.0) | 10.0 (7.0, 14.0) | 10.0 (7.0, 14.0) | <0.001 |
| No post-procedure aortic paravalvular regurgitation, (%) | 68.7 | 67.2 | 61.2 | <0.001 | 69.6 | 70.7 | 68.1 | <0.001 |
| Length of stay (days)** | 1.0 (1.0, 2.0) | 1.0 (1.0, | 1.0 (1.0, 3.0) | <0.001 | 1.0 (1.0, | 1.0 (1.0, | 1.0 (1.0, | <0.001 |
|  |  | 2.0) |  |  | 3.0) | 2.0) | 2.0) |  |
| *Hospital characteristics* |  |  |  |  |  |  |  |  |
| Bed size, (median, IQR) | 344 ( 250, 425) | 389 ( 288, 519) | 539 ( 380, 753) | <0.001 |  |  |  |  |
| Region, %: |  |  |  | 0.001 |  |  |  |  |
| • South<br>• Midwest<br>• West<br>• Northeast | • 39.9<br>• 25.1<br>• 20.9<br>• 12.9 | • 39.5<br>• 26.1<br>• 23.8<br>• 10.0 | • 33.3<br>• 22.7<br>• 19.7<br>• 23.9 |  |  |  |  |  |
| Type of hospital, % |  |  |  | <0.001 |  |  |  |  |
| • Rural<br>• Suburban<br>• Urban | • 18.6<br>• 32.3<br>• 49.0 | • 11.5<br>• 30.7<br>• 57.9 | • 6.1<br>• 28.4<br>• 65.5 |  |  |  |  |  |
| Teaching hospital, % | 51.7 | 51.0 | 70.5 | <0.001 |  |  |  |  |

\* STS PROM for isolated surgical aortic valve replacement (for TAVR patients)
eGFR based on MDRD equation
\*\* for patients discharged alive

At 1 year, the overall crude rates of all-cause mortality, stroke, and all-cause readmissions were 11.3%, 3.7%, and 37.9%, respectively, while median home-time was 363 days. (**Table 2**)

**Table 2.** Risk-adjusted rates and outcomes by annualized hospital and operator TAVR volume tertiles.

|  | Hospital |  |  |  | Operator |  |  |  |
| --- | --- | --- | --- | --- | --- | --- | --- | --- |
|  | **Tertile 1**<br>**(≤ 52/year)**<br>**(N = 23,645;**<br>**Hospitals =**<br>**263)** | **Tertile 2**<br>**(53-**<br>**101/year)**<br>**(N = 56,791;**<br>**Hospitals =**<br>**261)** | **Tertile 3**<br>**(≥102/year)**<br>**(N =**<br>**134,899;**<br>**Hospitals =**<br>**264)** | **Adjusted p-**<br>**value** | **Tertile 1**<br>**(≤ 11/year)**<br>**(N = 11822;**<br>**Operators =**<br>**1030)** | **Tertile 2**<br>**(12-24/year)**<br>**(N = 32000;**<br>**Operators =**<br>**1256)** | **Tertile 3**<br>**(≥25/year)**<br>**(N = 17113;**<br>**Operators =**<br>**1158)** | **Adjusted p-**<br>**value** |
| 1-year all-cause mortality, % | 9.5<br>OR: 1.10<br>(95% CI 1.05 – 1.16) | 9.0<br>OR: 1.05<br>(95% CI 1.00 – 1.09) | 8.7 | *0.0018* | 9.2<br>OR: 1.06<br>(95% CI 1.00 – 1.13) | 8.8<br>OR: 1.01<br>(95% CI 0.97 – 1.06) | 8.7 | *0.17* |
| 1-year stroke, % | 3.7<br>OR: 1.10<br>(95% CI 1.01 – 1.19) | 3.4<br>OR: 1.02<br>(95% CI 0.96 – 1.08) | 3.4 | *0.0085* | 3.9<br>OR: 1.16<br>(95% CI 1.05 – 1.28) | 3.4<br>OR: 1.03<br>(95% CI 0.96 – 1.1) | 3.4 | *0.011* |
| 1-year all-cause mortality or stroke, % | 12.3<br>OR: 1.10<br>(95% CI 1.05 – 1.15) | 11.7<br>OR: 1.03<br>(95% CI 0.99 – 1.07) | 11.4 | *0.0003* | 12.3<br>OR: 1.09<br>(95% CI 1.03 – 1.15) | 11.6<br>OR: 1.02<br>(95% CI 0.98 – 1.06) | 11.4 | *0.015* |
| 1-year readmission | 38.0<br>OR: 1.05<br>(95% CI 1.00 | 37.2<br>OR: 1.01<br>(95% CI 0.98 | 36.9 | *0.035* | 37.5<br>OR: 1.02<br>(95% CI 0.98 | 37.2<br>OR: 1.01<br>(95% CI 0.98 | 37.0 | *0.77* |
|  | – 1.09) | – 1.05) |  |  | – 1.07) | – 1.04) |  |  |

CI = confidence intervals; OR = odds ratio

Association between annual hospital TAVR volumes and outcomes (**Figure 2**, **Table 2.)**:

**Figure 2:**
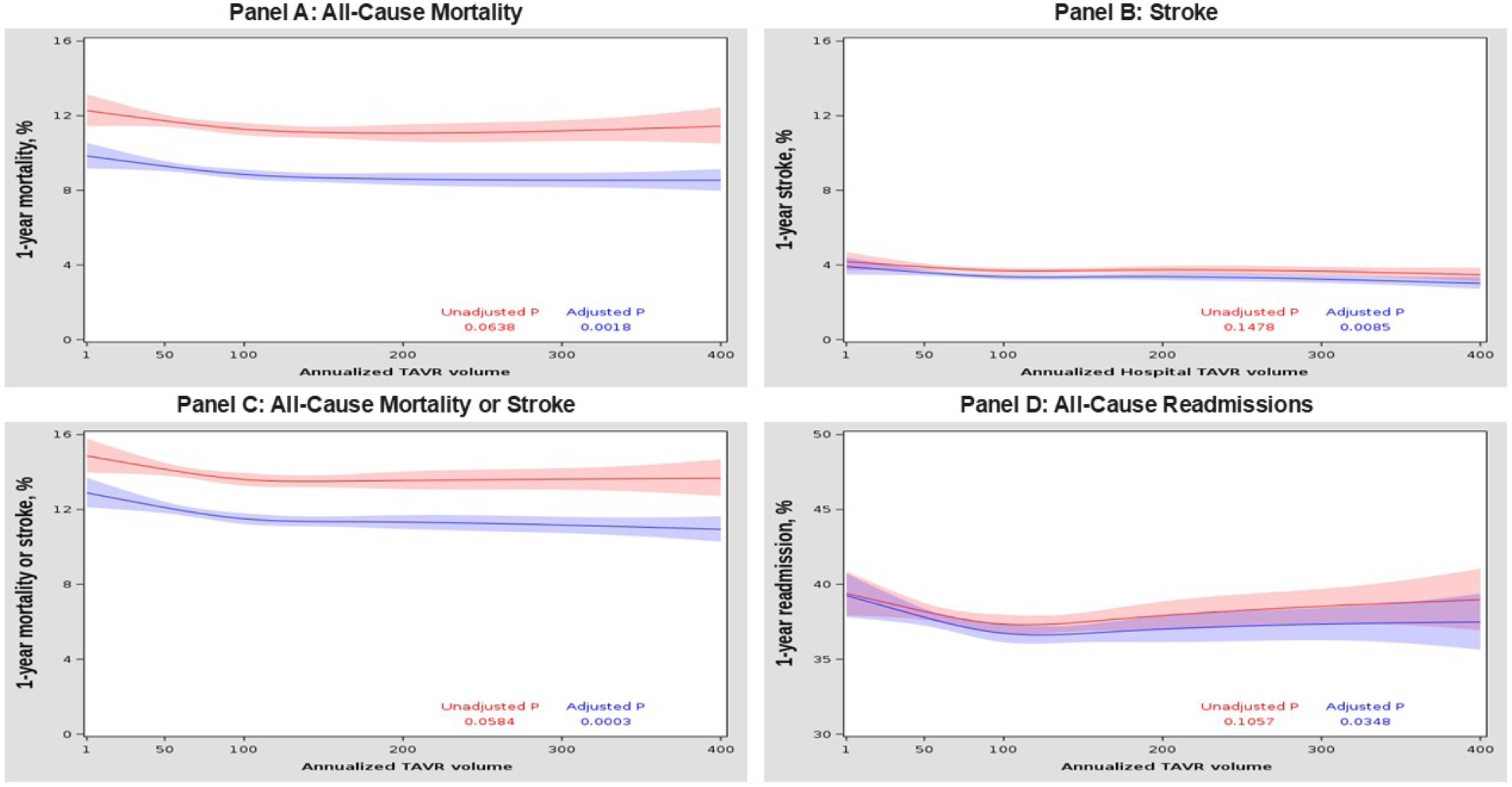
Relationship between annual hospital volumes and 1-year all-cause mortality (panel A), stroke (panel B), mortality or stroke (panel C) and readmissions (panel D).

Hospitals were categorized by annual TAVR volumes into low (≤52/year), medium (53-101/year), and high (≥ 102/year) tertiles. Compared with high-volume hospitals, low-volume hospitals had higher risk-adjusted rates of 1-year all-cause mortality (12.0% vs. 11.2%; OR=1.10 ; 95% CI 1.05-1.16; adjusted p-value=0.0018; NNH=125), 1-year stroke (4.0% vs. 3.7%, OR=1.10, 95% CI 1.01–1.19; adjusted p-value=0.0085; NNH = 334), composite of 1-year all-cause mortality or stroke (14.5% vs. 13.6%, OR=1.10, 95% CI 1.05-1.15, adjusted p-value=0.0003; NNH = 112), 1-year all-cause readmission rates (38.5% vs. 38.0%, OR=1.05, 95% CI 1.0-1.09; adjusted p-value=0.035; NNH = 91) and 1-year home time (335.2 vs. 333 days, IRR 0.99, 95% CI 0.99-1.00, adjusted p-value=0.02). One-year mortality, stroke and all-cause mortality or stroke increased progressively across TAVR volume tertiles, with the curves for low-volume hospitals appearing to diverge from the other two categories around 3 to 6 months (**Figure 3**).

**Figure 3:**
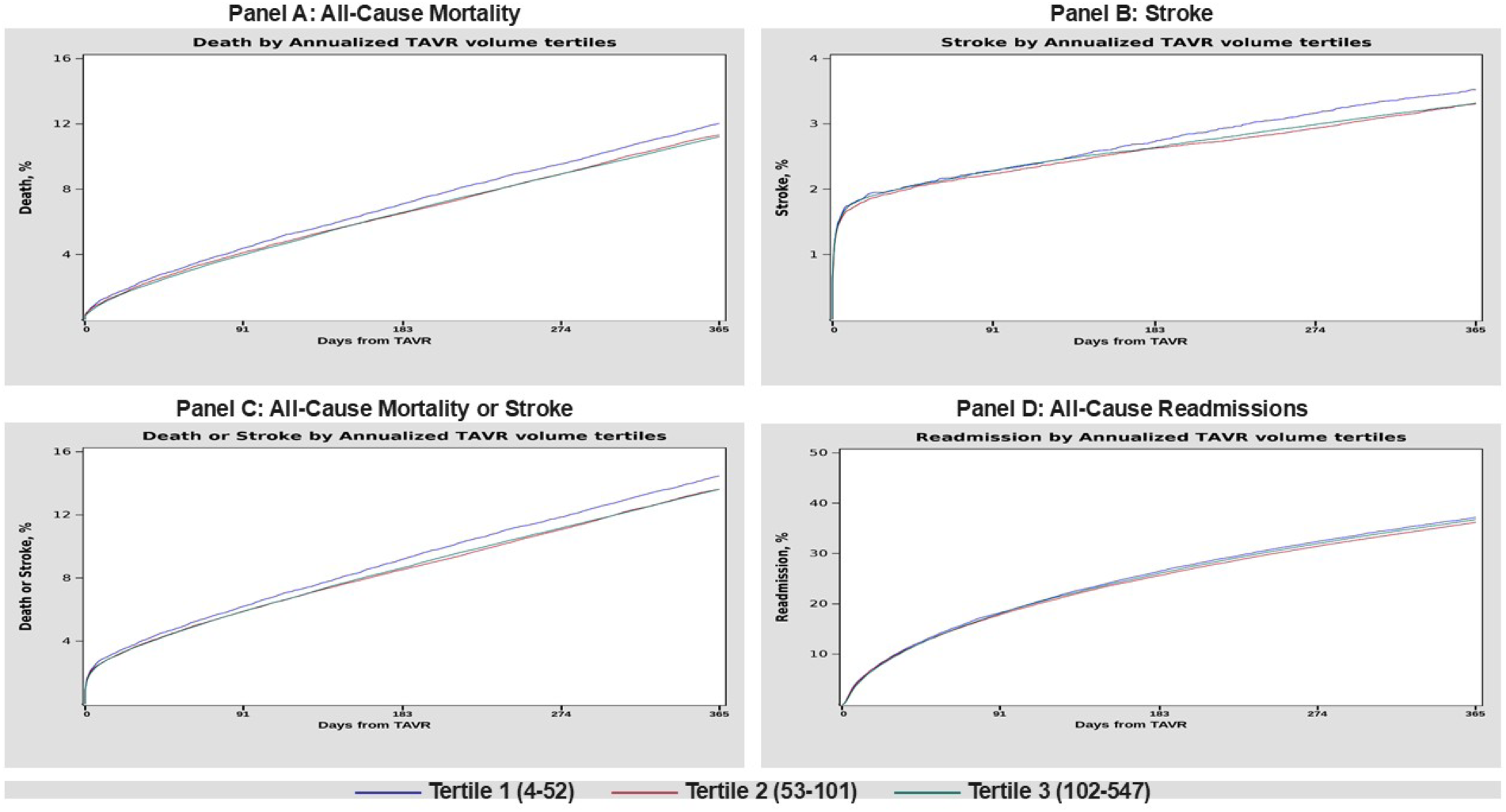
Relationship between annual hospital volume tertiles and 1-year all-cause mortality (panel A), stroke (panel B), mortality or stroke (panel C) and readmissions (panel D).

Association between annual operator TAVR volumes and outcomes (**Figure 4**, **Table 2):**

**Figure 4:**
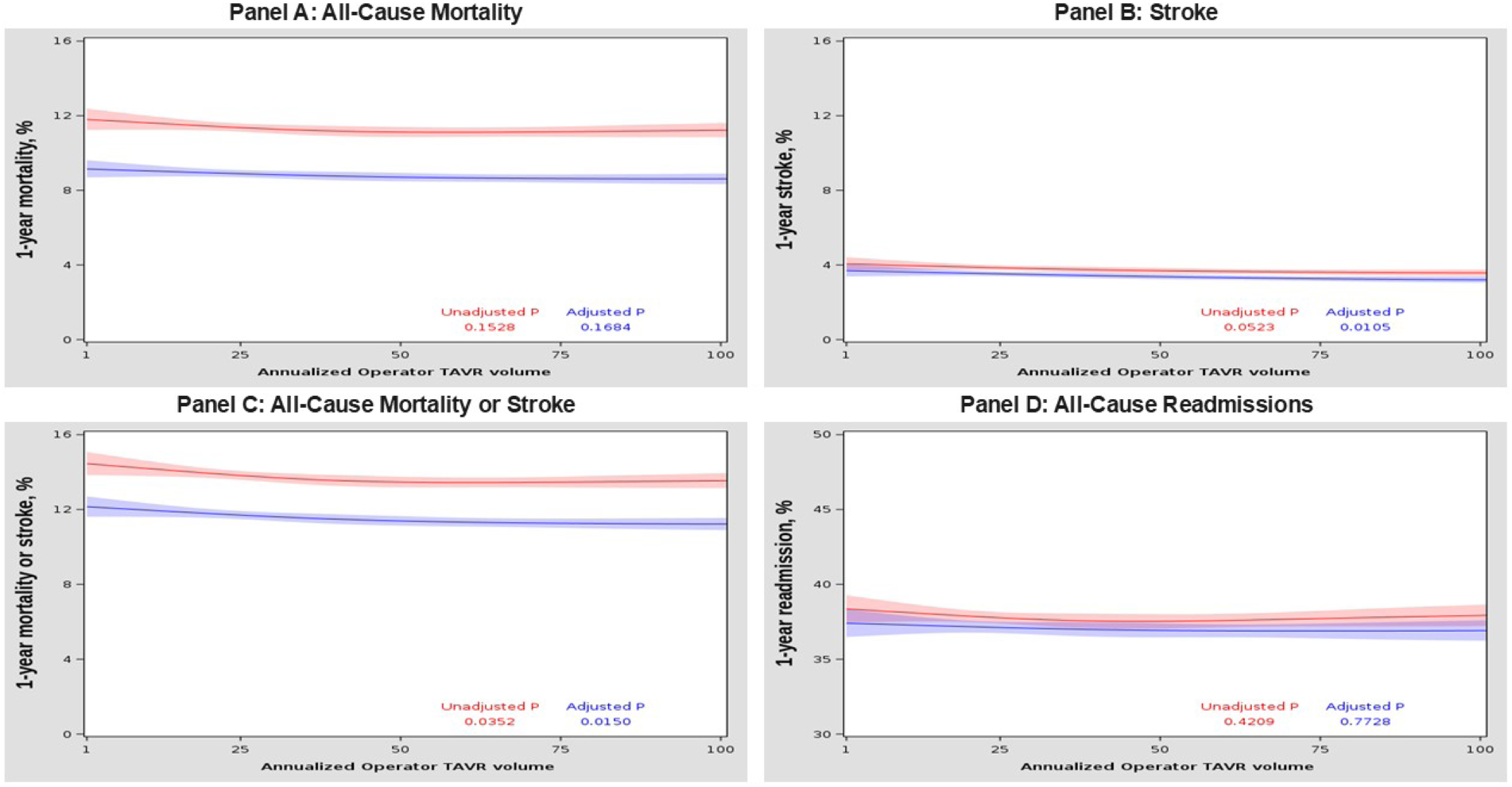
Relationship between annual operator volumes and 1-year all-cause mortality (panel A), stroke (panel B), mortality or stroke (panel C) and readmissions (panel D).

Operators were categorized by annual TAVR volumes into low (≤11/year), medium (12-24/year), and high (≥25/year) tertiles. Compared with high-volume operators, low-volume operators did not have stastically significant different risk-adjusted rates of 1-year all-cause mortality (11.9% vs. 11.3%, OR=1.06 ; 95% CI 1.00-1.13; adjusted p-value=0.17) or 1-year home-time (334.9 vs. 333.0 days, IRR = 0.99, 95% CI 0.99-1.00. adjusted p-value=0.25), but higher rates of 1-year stroke (4.2% vs. 3.7%, OR=1.16, 95% CI 1.05–1.28; adjusted p-value=0.011; NNH = 200) and 1-year all-cause mortality or stroke (14.6% vs. 13.6%, OR=1.09, 95% CI 1.03-1.15, adjusted p-value=0.015; NNH = 112). There was no significant difference in 1-year readmission rates (38.4% vs. 37.9%, OR=1.02, 95% CI 0.98-1.07; adjusted p-value=0.77). One-year mortality, stroke and composite of all-cause mortality or stroke increased progressively across follow-up in all annualized TAVR volume tertiles, with the curves for low-volume operators appearing to diverge early (∼30 days) from the other two categories for stroke and stroke and all-cause mortality, and around 3 to 6 months for all-cause mortality (**Figure 5**).

**Figure 5:**
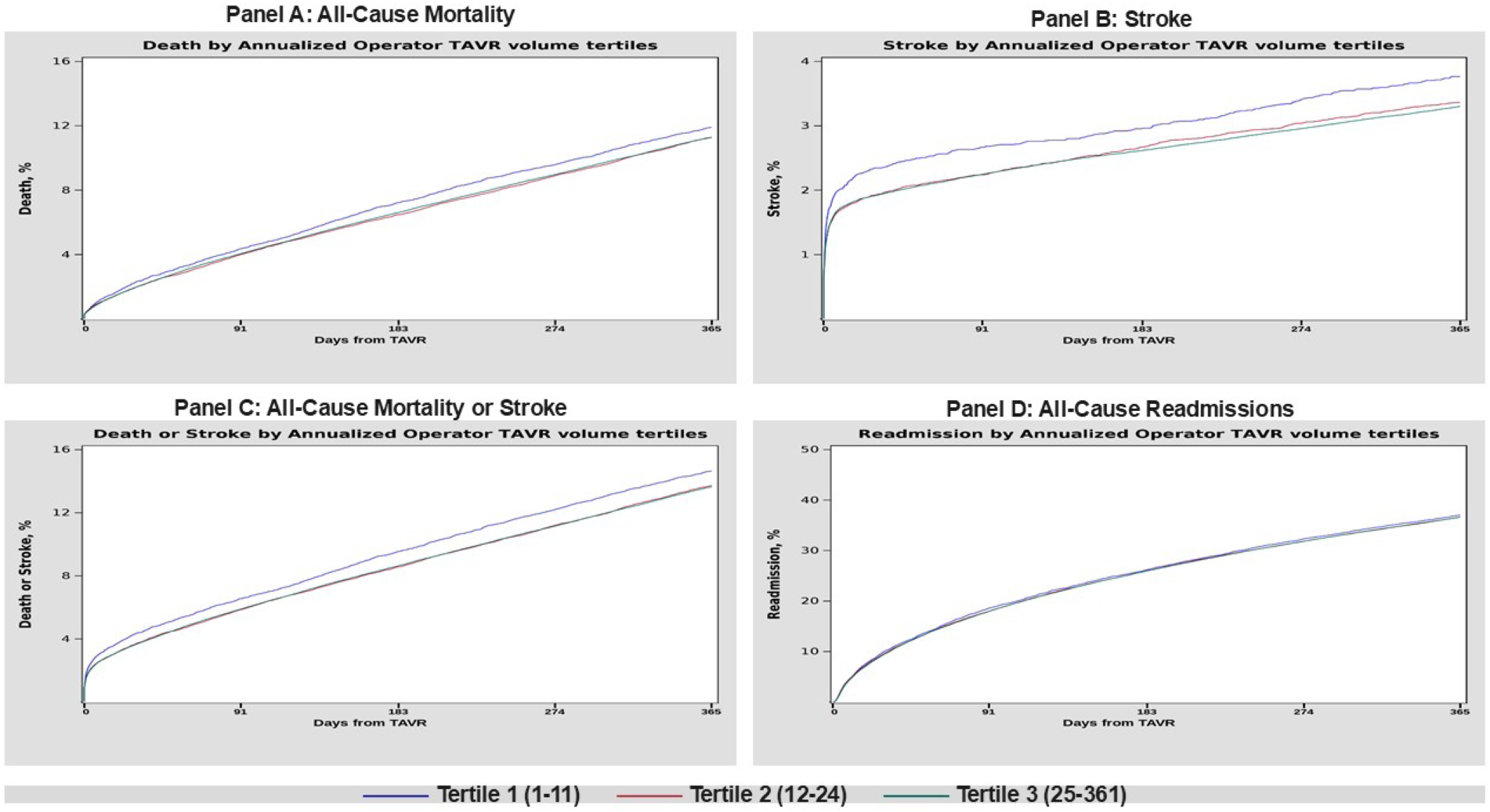
Relationship between annual operator volume tertiles and 1-year all-cause mortality (panel A), stroke (panel B), mortality or stroke (panel C) and readmissions (panel D).

### Interaction between hospital and operator volumes for TAVR

The interaction p-values between hospital and operator volumes were not significant for all 4 endpoints – all-cause mortality (p=0.87), stroke (p=0.80), all-cause mortality or stroke (p=0.90) and readmission (p=0.42).

## Discussion

Our study leveraged data from the STS/ACC TVT-R linked to CMS administrative claims to evaluate 1-year outcomes among >215,000 patients undergoing TAVR at 788 sites by 3,444 TAVR operators between 2020 and 2022. This represents one of the largest studies evaluating one-year outcomes in this population and the relationship between annualized hospital and operator volumes and clinical outcomes. We made several key observations. Despite treating patients with lower baseline risk and fewer comorbidities, low-volume hospitals and operators experienced higher rates of adverse 1-year outcomes, consistent with an inverse association between procedural volume and outcomes. Compared with high-volume hospitals (≥102 cases/year), low-volume hospitals (≤52 cases/year; on average ≤ 1 case/week) had approximately 5%-10% higher odds of mortality, stroke, or the composite of mortality or stroke at 1 year, with event curves separating at approximately 3–6 months. Similarly, compared with high-volume operators (≥25 cases/year), low-volume operators (≤11 cases/year; on average, < 1 case/month) had 5%-15% higher odds of stroke and the composite of all-cause mortality or stroke, with earlier separation of event curves (∼30 days). Hospital readmissions were inversely associated with hospital volume, but not operator volume. There was no significant interaction between hospital and operator volume for any of the 4 endpoints, suggesting that the associations between hospital and operator volume and 1-year outcomes were independent of each other.

Volume thresholds have long played an important role as surrogates for procedural quality. (17–23) For most procedures, there is initially a steeper volume-outcomes association, reflecting an early learning curve. As the procedure matures, operators gain experience, procedural and device refinements occur, and the eligible patient population expands to include lower risk patients. Consequently, the volume-outcomes association may begin to plateau and become apparent only for outcomes with higher event rates (e.g., composite endpoints) or over longer durations of follow-up. In fact, in an ideal scenario, procedural volume would no longer serve as a surrogate for quality; instead, the focus would shift toward direct assessment of quality benchmarks to facilitate continuous quality improvement.(24)

This has indeed been true for the TAVR experience as well. Introduced in the US in 2011 for patients at high and extreme surgical risk, TAVR initially represented a high-complexity, high-risk procedure, with in-hospital mortality of 4-6% and a median length of stay of 6 days. (7,25) Early volume-outcomes analyses from TVT-R (2011-2015) from 395 hospitals demonstrated a steep association between procedural volume and outcomes, with absolute differences in in-hospital mortality exceeding 1% across institutional volume strata.(7) A similar operator-level association was noted in a subsequent analysis as well. (8) Over the subsequent decade, marked improvements in patient selection, device technology, and procedural standardization have transformed TAVR into a highly streamlined intervention. In contemporary practice, the procedure can be performed with risk comparable to complex PCI, with 30-day mortality of approximately 2% and a median post-procedural length of stay of 1 day at most centers. In two recent analyses of patients in the contemporary low-risk era, we observed a plateauing of the TAVR volume-outcomes relationship at 30 days, particularly at the hospital level,(9) suggesting that the incremental benefit of increasing institutional volumes may have diminished as TAVR has become more standardized and widely adopted across centers. Despite TAVR now being performed at more than 800 sites across the United States, a modest but persistent association between operator volume and outcomes remained evident, underscoring the additional importance of procedural expertise even as the overall TAVR ecosystem has matured.(10) Importantly, the present findings suggest a predominantly “low versus not low” volume-outcomes relationship, with minimal additional separation between mid- and high-volume centers. The operator level volume thresholds identified in the present analysis (≤11, 12-24, and ≥25 cases/year) were similar, albeit slightly lower, than those reported in our prior analysis (≤14, 15–36, ≥37 cases/year), likely reflecting restriction of the present cohort to Medicare eligible patients.(10)

Notwithstanding the possibility of performance outliers independent of procedural volume and variability within volume strata, the present findings suggest that volume thresholds remain a relevant and potentially important policy lever for maintaining procedural safety and optimizing outcomes. In particular, the persistence of a volume-outcomes relationship in the contemporary era supports continued consideration of minimum volume standards within future regulatory and accreditation frameworks for TAVR programs and operators. While the operator-level association was evident at both 30 days and 1 year, the hospital-level association appeared to emerge primarily over longer-term follow-up, suggesting that institutional factors may exert a more cumulative influence on outcomes beyond the periprocedural period. These findings are particularly relevant given that the recently proposed TAVR NCD places primary emphasis on operator competency and procedural volumes,(26) yet our results suggest that hospital-level experience and systems of care may independently contribute to longer-term outcomes (death, stroke, readmissions) and therefore warrant consideration within future regulatory frameworks.

Any such requirements, however, must be balanced against potential effects on access to care.(27,28) If a causal relationship is assumed, our NNH estimates suggest that approximately 112 patients would need to be treated at high-volume rather than lower-volume centers to prevent one death or stroke at 1 year. Applied to the study population, this effect size corresponds to approximately 211 excess deaths or strokes at 1 year among patients treated at low-volume hospitals during the 2-year study period. These estimates provide a clinically interpretable measure of the tradeoff between procedural quality and access. Although TAVR operators and hospitals were predominantly located in urban areas, the relatively greater representation of minorities and rural hospitals and operators among lower-volume programs suggests that these centers may play an important role in expanding access to TAVR. This tradeoff is likely to assume greater importance over time as the prevalence of AS continues to rise with population aging. In the absence of effective therapies to prevent or delay disease progression, increasing numbers of patients will require valve intervention.(3,21) Moreover, structural valve degeneration following both surgical and transcatheter bioprosthetic valve replacement is generating an additional and rapidly growing population of patients who may require repeat treatment,(29,30) further amplifying the demand for TAVR services.

An important observation was that patients undergoing TAVR at low-volume hospitals and by low-volume operators were, on average, lower risk, yet were more likely to undergo general anesthesia and experienced longer procedure times. In our prior analysis, we observed similar differences in process-of-care measures between low- and higher-volume operators, including greater contrast utilization and worse post-procedural mean aortic valve gradients and higher rates of paravalvular regurgitation compared with high-volume operators.(10) Others have reported an inverse association between hospital TAVR volumes and failure to rescue from major in-hospital complications.(31) Collectively, these findings suggest plausible mechanistic pathways through which lower procedural volume may translate into worse longer-term outcomes, despite treatment of ostensibly lower-risk patients, as well as possible interventions to improve them. This is an important consideration for access discussions as well,(32) and supports the robustness and clinical relevance of the volume-outcome association.

In prior analyses comparing administrative claims-based outcomes with clinical events committee-adjudicated endpoints, claims-based ascertainment of mortality has demonstrated very high accuracy.(33) Despite a lower reported sensitivity for stroke ascertainment using claims-based data,(34,35) the stroke rates observed in the current analysis appear similar to prior reports based on adjudicated clinical outcomes.(36,37) We focused on outcomes through 1 year, as longer-term events are increasingly influenced by post-procedural management (including by other clinicians and hospitals) and patient-level factors rather than the quality of the index TAVR procedure. The differing temporal patterns observed for operator- and hospital-level associations are noteworthy. Divergence in operator-level outcomes was evident by 30 days, a timeframe closely linked to procedural performance and technical expertise. In contrast, separation of hospital-level outcome curves emerged after 3-6 months, suggesting contributions from factors beyond the procedure itself, including early post-procedural care, discharge planning, institutional infrastructure, and longitudinal follow-up processes.

We acknowledge several limitations of this analysis. First, as a retrospective observational study, the findings remain susceptible to residual and unmeasured confounding despite comprehensive risk adjustment, particularly given that the study period encompassed the COVID-19 pandemic. Our linkage relied on deterministic matching using indirect identifiers; and therefore, linkage failures and mislinks are possible and may introduce selection or classification bias. Because TVT-R excludes trial procedures, operator and hospital volumes may have been underestimated at participating centers. CMS requires TAVR procedures in the U.S. to be jointly performed by an interventional cardiologist and a cardiac surgeon, making primary operator designation and attribution of outcomes challenging. Consistent with prior studies, we designated the operator with the greatest cumulative TAVR experience as the primary operator.(10) Data on dedicated structural fellowship training were unavailable, and lifetime operator experience could only be indirectly measured.

In conclusion, our analysis of a large, contemporary CMS-linked national TAVR registry demonstrates an inverse association between annual hospital and operator procedural volumes and 1-year clinical outcomes. Annual hospital volumes ≤52 and operator volumes ≤11 cases were independently associated with worse outcomes compared with higher volume hospitals and operators, respectively. These findings may inform future quality and policy initiatives and help identify potential targets for quality improvement.

## Data Availability

Data from the present study are not publicly available, however, requests for data access may be made to the STS/ACC TVT Registry at for consideration.

## References

1. Carroll JD, Mack MJ, Vemulapalli S et al. STS-ACC TVT Registry of Transcatheter Aortic Valve Replacement. J Am Coll Cardiol 2020;76:2492–2516.

2. Otto CM, Kumbhani DJ, Alexander KP et al. 2017 ACC Expert Consensus Decision Pathway for Transcatheter Aortic Valve Replacement in the Management of Adults With Aortic Stenosis: A Report of the American College of Cardiology Task Force on Clinical Expert Consensus Documents. J Am Coll Cardiol 2017;69:1313–1346.

3. Kumbhani DJ, Pandey A. From Passive to Proactive: A Call to Action on Valve Disease Screening. Circulation 2025;151:1508–1511.

4. Sukul D, Allen J, Kumbhani DJ. Volume Considerations for Transcatheter Aortic Valve Replacement in Medicare’s National Coverage Determination. Circ Cardiovasc Qual Outcomes 2019;12:e005216.

5. https://www.cms.gov/medicare-coverage-database/view/ncd.aspx?NCDId=355. Accessed June 24, 2026.

6. https://www.cms.gov/medicare-coverage-database/view/nca.aspx?ncaid=293. Accessed June 24, 2026.

7. Carroll JD, Vemulapalli S, Dai D et al. Procedural Experience for Transcatheter Aortic Valve Replacement and Relation to Outcomes: The STS/ACC TVT Registry. J Am Coll Cardiol 2017;70:29–41.

8. Vemulapalli S, Carroll JD, Mack MJ et al. Procedural Volume and Outcomes for Transcatheter Aortic-Valve Replacement. N Engl J Med 2019;380:2541–2550.

9. Kumbhani DJ, Manandhar P, Bavry AA et al. National Variation in Hospital MTEER Outcomes and Correlation With TAVR Outcomes: STS/ACC TVT Registry Analysis. JACC Cardiovasc Interv 2024;17:505–515.

10. Kumbhani DJ, Girotra S, Dong H et al. Contemporary Operator Procedural Volumes and Outcomes for TAVR and MTEER in the US. JAMA Cardiol 2026.

11. Carroll JD, Edwards FH, Marinac-Dabic D et al. The STS-ACC transcatheter valve therapy national registry: a new partnership and infrastructure for the introduction and surveillance of medical devices and therapies. J Am Coll Cardiol 2013;62:1026–34.

12. Hammill BG, Hernandez AF, Peterson ED, Fonarow GC, Schulman KA, Curtis LH. Linking inpatient clinical registry data to Medicare claims data using indirect identifiers. Am Heart J 2009;157:995–1000.

13. Brennan JM, Thomas L, Cohen DJ et al. Transcatheter Versus Surgical Aortic Valve Replacement: Propensity-Matched Comparison. J Am Coll Cardiol 2017;70:439–450.

14. Vemulapalli S, Dai D, Hammill BG et al. Hospital Resource Utilization Before and After Transcatheter Aortic Valve Replacement: The STS/ACC TVT Registry. J Am Coll Cardiol 2019;73:1135–1146.

15. Columbo JA, Daya N, Colantonio LD et al. Derivation and Validation of ICD-10 Codes for Identifying Incident Stroke. JAMA Neurol 2024;81:875–881.

16. Mentias A, Keshvani N, Desai MY, et al. Risk-Adjusted, 30-Day Home Time After Transcatheter Aortic Valve Replacement as a Hospital-Level Performance Metric. J Am Coll Cardiol 2022;79:132–144.

17. Khera R, Pandey A, Koshy T et al. Role of Hospital Volumes in Identifying Low-Performing and High-Performing Aortic and Mitral Valve Surgical Centers in the United States. JAMA Cardiol 2017;2:1322–1331.

18. Kumbhani DJ, Bittl JA. Much Ado About Nothing? The Relationship of Institutional Percutaneous Coronary Intervention Volume to Mortality. Circ Cardiovasc Qual Outcomes 2017;10.

19. Kumbhani DJ, Fonarow GC, Heidenreich PA et al. Association Between Hospital Volume, Processes of Care, and Outcomes in Patients Admitted With Heart Failure: Insights From Get With The Guidelines-Heart Failure. Circulation 2018;137:1661–1670.

20. Kumbhani DJ, Nallamothu BK. PCI Volume Benchmarks: Still Adequate for Quality Assessment in 2017? J Am Coll Cardiol 2017;69:2925–2928.

21. Kumbhani DJ, Peterson ED. Sequential Evolution of Quality Assessment for Aortic Valvular Heart Interventions. Circulation 2021;144:195–198.

22. Kumbhani DJ, Cannon CP, Fonarow GC et al. Association of hospital primary angioplasty volume in ST-segment elevation myocardial infarction with quality and outcomes. JAMA 2009;302:2207–13.

23. Elbadawi A, Mohamed A, Sedhom R et al. Clinical Outcomes in Relation to Total Hospital Surgical and Transcatheter Aortic Valve Replacement Volumes. J Am Heart Assoc 2024;13:e035719.

24. Bavaria JE, Tommaso CL, Brindis RG et al. 2018 AATS/ACC/SCAI/STS Expert Consensus Systems of Care Document: Operator and Institutional Recommendations and Requirements for Transcatheter Aortic Valve Replacement: A Joint Report of the American Association for Thoracic Surgery, American College of Cardiology, Society for Cardiovascular Angiography and Interventions, and Society of Thoracic Surgeons. J Am Coll Cardiol 2019;73:340–374.

25. Grover FL, Vemulapalli S, Carroll JD et al. 2016 Annual Report of The Society of Thoracic Surgeons/American College of Cardiology Transcatheter Valve Therapy Registry. J Am Coll Cardiol 2017;69:1215–1230.

26. https://www.cms.gov/medicare-coverage-database/view/nca.aspx?ncaid=321.

27. Nelson AJ, Wegermann ZK, Gallup D et al. Modeling the Association of Volume vs Composite Outcome Thresholds With Outcomes and Access to Transcatheter Aortic Valve Implantation in the US. JAMA Cardiol 2023;8:492–502.

28. Marquis-Gravel G, Stebbins A, Kosinski AS et al. Geographic Access to Transcatheter Aortic Valve Replacement Centers in the United States: Insights From the Society of Thoracic Surgeons/American College of Cardiology Transcatheter Valve Therapy Registry. JAMA Cardiol 2020;5:1006–1010.

29. Kumbhani DJ, Chhatriwalla AK. VIVID Insights: Tips for Successful Valve-in-Valve TAVR in Stented and Stentless Surgical Valves. JACC Cardiovasc Interv 2019;12:1264–1267.

30. Bavry AA, Kumbhani DJ. As Patients Live Longer, Are We on the Cusp of a New Valve Epidemic? J Am Coll Cardiol 2021;77:15–17.

31. Sreenivasan J, Vemulapalli S, Kosinski A et al. Institutional Volume and Failure to Rescue in Transcatheter Aortic Valve Replacement: Insights From the STS/ACC TVT Registry. J Am Coll Cardiol 2024;84:B367.

32. Mack MJ, Holmes DR, Jr. Rational dispersion for the introduction of transcatheter valve therapy. JAMA 2011;306:2149–50.

33. Butala NM, Strom JB, Faridi KF et al. Validation of Administrative Claims to Ascertain Outcomes in Pivotal Trials of Transcatheter Aortic Valve Replacement. JACC Cardiovasc Interv 2020;13:1777–1785.

34. Strom JB, Zhao Y, Faridi KF et al. Comparison of Clinical Trials and Administrative Claims to Identify Stroke Among Patients Undergoing Aortic Valve Replacement: Findings From the EXTEND Study. Circ Cardiovasc Interv 2019;12:e008231.

35. Kumbhani DJ, Welt FG. Supplementing Randomized Trial Data to Answer a Real-World Question: Discharge to Home Status as a Heuristic for Stroke Severity After TAVR. Circ Cardiovasc Interv 2024;17:e014374.

36. Mack MJ, Leon MB, Thourani VH et al. Transcatheter Aortic-Valve Replacement with a Balloon-Expandable Valve in Low-Risk Patients. N Engl J Med 2019;380:1695–1705.

37. van Nieuwkerk AC, Aarts HM, Hemelrijk KI et al. Cerebrovascular Events in Patients Undergoing Transfemoral Transcatheter Aortic Valve Implantation: A Pooled Patient-Level Study. J Am Heart Assoc 2024;13:e032901.

