## supplemental figure and methods for "Hospital and operator procedural volumes and one-year outcomes for TAVR in the United States: A STS/ACC TVT Registry analysis"

### Supplemental material

Supplemental Figure: CONSORT flow diagram of patient inclusion for TAVR

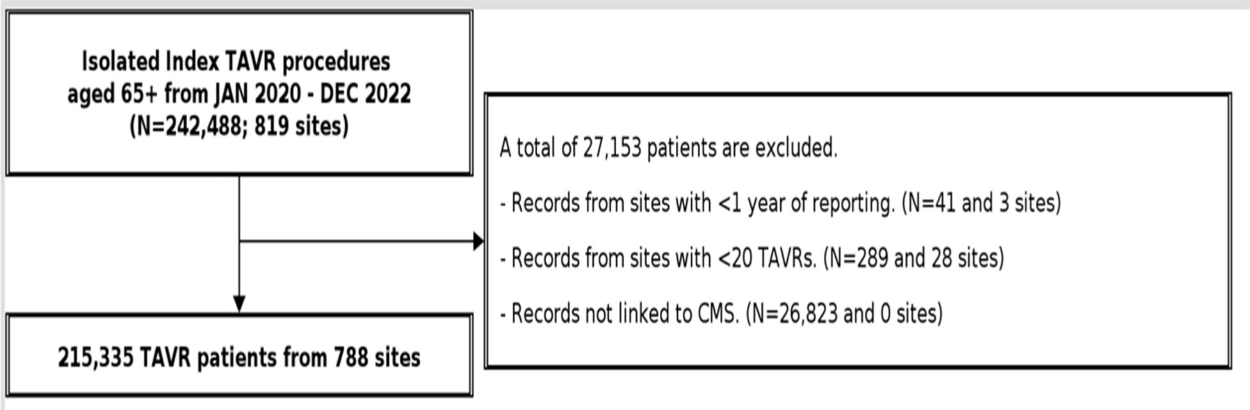

Supplemental Methods

*Variable adjustment list for TAVR*

| | | |
|---|---|---|
| Age | Sex | Race/Hispanic Origin |
| Sex-specific BSA | LVEF | HGB |
| Platelet count | GFR | Dialysis |
| Left main >=50% | Proximal LAD >= 70% | Prior MI |
| Endocarditis | Prior Stroke/TIA | Carotid Stenosis |
| Prior PAD | Current Smoker | Diabetes |
| Nyha Class 4 | Atrial fibrillation/flutter | Conduction Defect |
| Severe Lung Disease | Home Oxygen | Hostile Chest |
| Porcelain Aorta | Pacemaker | Previous PCI |
| Prior CABG | Previous cardiac surgeries | Prior aortic valve procedure |
| Prior non-aortic valve procedure | | |

<u>Supplemental Methods</u>

*Variables with > 2% missing data:*

- Left main stenosis: 5.4%
- Proximal LAD stenosis: 5.6%
